## Supplementary material for "Triaging and Referring In Adjacent General and Emergency Departments (the TRIAGE trial): a cluster randomised controlled trial": CONSORT Check list

**Table 1: CONSORT 2010 checklist of information to include when reporting a cluster randomised trial**

| Section/Topic | Item No | Standard Checklist item | Extension for cluster designs | Page No * |
| --- | --- | --- | --- | --- |
| Title and abstract | | | |  |
|  | 1a | Identification as a randomised trial in the title | Identification as a cluster randomised trial in the title | 1 |
|  | 1b | Structured summary of trial design, methods, results, and conclusions (for specific guidance see CONSORT for abstracts)^[1],^^[2]^ | See table 2 | see below |
| Introduction | | | |  |
| Background and objectives | 2a | Scientific background and explanation of rationale | Rationale for using a cluster design | 4 |
|  | 2b | Specific objectives or hypotheses | Whether objectives pertain to the the cluster level, the individual participant level or both | 4 |
| Methods | | | |  |
| Trial design | 3a | Description of trial design (such as parallel, factorial) including allocation ratio | Definition of cluster and description of how the design features apply to the clusters | 5 |
|  | 3b | Important changes to methods after trial commencement (such as eligibility criteria), with reasons |  | 6 (refers to supplementary material) |
| Participants | 4a | Eligibility criteria for participants | Eligibility criteria for clusters | 5-6 |
|  | 4b | Settings and locations where the data were collected |  | 5 |
| Interventions | 5 | The interventions for each group with sufficient details to allow replication, including how and when they were actually administered | Whether interventions pertain to the cluster level, the individual participant level or both | 7 |
| Outcomes | 6a | Completely defined pre-specified primary and secondary outcome measures, including how and when they were assessed | Whether outcome measures pertain to the cluster level, the individual participant level or both | 7-8 |
|  | 6b | Any changes to trial outcomes after the trial commenced, with reasons |  | 6 (refers to supplementary material) |
| Sample size | 7a | How sample size was determined | Method of calculation, number of clusters(s) (and whether equal or unequal cluster sizes are assumed), cluster size, a coefficient of intracluster correlation (ICC or *k*), and an indication of its uncertainty | 8 |
|  | 7b | When applicable, explanation of any interim analyses and stopping guidelines |  | 10 |
| Randomisation: | | | |  |
| Sequence generation | 8a | Method used to generate the random allocation sequence |  | 8 |
|  | 8b | Type of randomisation; details of any restriction (such as blocking and block size) | Details of stratification or matching if used | 8 |
| Allocation concealment mechanism | 9 | Mechanism used to implement the random allocation sequence (such as sequentially numbered containers), describing any steps taken to conceal the sequence until interventions were assigned | Specification that allocation was based on clusters rather than individuals and whether allocation concealment (if any) was at the cluster level, the individual participant level or both | 8 |
| Implementation | 10 | Who generated the random allocation sequence, who enrolled participants, and who assigned participants to interventions | Replace by 10a, 10b and 10c | 8 |
|  | 10a |  | Who generated the random allocation sequence, who enrolled clusters, and who assigned clusters to interventions | 8 |
|  | 10b |  | Mechanism by which individual participants were included in clusters for the purposes of the trial (such as complete enumeration, random sampling) | N/A |
|  | 10c |  | From whom consent was sought (representatives of the cluster, or individual cluster members, or both), and whether consent was sought before or after randomisation | 10 |
| Blinding | 11a | If done, who was blinded after assignment to interventions (for example, participants, care providers, those assessing outcomes) and how |  | N/A |
|  | 11b | If relevant, description of the similarity of interventions |  | N/A |
| Statistical methods | 12a | Statistical methods used to compare groups for primary and secondary outcomes | How clustering was taken into account | 10-11 and the Statistical Analysis Plan |
|  | 12b | Methods for additional analyses, such as subgroup analyses and adjusted analyses |  | N/A |
| Results | | | |  |
| Participant flow (a diagram is strongly recommended) | 13a | For each group, the numbers of participants who were randomly assigned, received intended treatment, and were analysed for the primary outcome | For each group, the numbers of clusters that were randomly assigned, received intended treatment, and were analysed for the primary outcome | Figure 2 |
|  | 13b | For each group, losses and exclusions after randomisation, together with reasons | For each group, losses and exclusions for both clusters and individual cluster members | Figure 2 |
| Recruitment | 14a | Dates defining the periods of recruitment and follow-up |  | 5 |
|  | 14b | Why the trial ended or was stopped |  | N/A |
| Baseline data | 15 | A table showing baseline demographic and clinical characteristics for each group | Baseline characteristics for the individual and cluster levels as applicable for each group | Table 1 |
| Numbers analysed | 16 | For each group, number of participants (denominator) included in each analysis and whether the analysis was by original assigned groups | For each group, number of clusters included in each analysis | Figure 2 |
| Outcomes and estimation | 17a | For each primary and secondary outcome, results for each group, and the estimated effect size and its precision (such as 95% confidence interval) | Results at the individual or cluster level as applicable and a coefficient of intracluster correlation (ICC or k) for each primary outcome | 13-19 |
|  | 17b | For binary outcomes, presentation of both absolute and relative effect sizes is recommended |  | 13-19 |
| Ancillary analyses | 18 | Results of any other analyses performed, including subgroup analyses and adjusted analyses, distinguishing pre-specified from exploratory |  | 18-19  Only the exploration of the primary ouctome after the trial was not pre-specified. |
| Harms | 19 | All important harms or unintended effects in each group (for specific guidance see CONSORT for harms^[3]^) |  | 16 (refers to the supplementary material) |
| Discussion | | | |  |
| Limitations | 20 | Trial limitations, addressing sources of potential bias, imprecision, and, if relevant, multiplicity of analyses |  | 19-20 |
| Generalisability | 21 | Generalisability (external validity, applicability) of the trial findings | Generalisability to clusters and/or individual participants (as relevant) | 19-20 |
| Interpretation | 22 | Interpretation consistent with results, balancing benefits and harms, and considering other relevant evidence |  | 20 |
| Other information | | |  |  |
| Registration | 23 | Registration number and name of trial registry |  | abstract |
| Protocol | 24 | Where the full trial protocol can be accessed, if available |  | 6 (refers to the Supplementary material) |
| Funding | 25 | Sources of funding and other support (such as supply of drugs), role of funders |  | Abstract and Source of support statement |

** Note: page numbers optional depending on journal requirements*

**Table 2:	Extension of CONSORT for abstracts**1**^,^**2 **to reports of cluster randomised trials (applicable to the abstract found in the main document)**

| **Item** | **Description** | **Reported on line number** |
| --- | --- | --- |
| Title | Identification of the study as randomized | 14 |
| Authors * | Contact details for the corresponding author | Title page |
| Trial design | Description of the trial design (e.g. parallel, cluster, non-inferiority) | 18 |
| Methods |  |  |
| Participants | Eligibility criteria for participants and the settings where the data were collected | 15-17 |
| Interventions | Interventions intended for each group | 19-21 |
| Objective | Specific objective or hypothesis | 15 |
| Outcome | Clearly defined primary outcome for this report | 22 |
| Randomization | How participants were allocated to interventions | 23-24 |
| Blinding (masking) | Whether or not participants, care givers, and those assessing the outcomes were blinded to group assignment | Not necessary |
| Results |  |  |
| Numbers randomized | Number of participants randomized to each group | 29-30 |
| Recruitment* | Trial status | N/A |
| Numbers analysed | Number of participants analysed in each group | 18-19 |
| Outcome | For the primary outcome, a result for each group and the estimated effect size and its precision | 29-31 |
| Harms | Important adverse events or side effects | Only 300 words allowed |
| Conclusions | General interpretation of the results | 37 |
| Trial registration | Registration number and name of trial register | 39 |
| Funding | Source of funding | 40 |

**this item is specific to conference abstracts*

1. Hopewell S, Clarke M, Moher D, Wager E, Middleton P, Altman DG, et al. CONSORT for reporting randomised trials in journal and conference abstracts. *Lancet* 2008, 371:281-283
2. Hopewell S, Clarke M, Moher D, Wager E, Middleton P, Altman DG at al (2008) CONSORT for reporting randomized controlled trials in journal and conference abstracts: explanation and elaboration. *PLoS Med* 5(1): e20
3. Ioannidis JP, Evans SJ, Gotzsche PC, O'Neill RT, Altman DG, Schulz K, Moher D. Better reporting of harms in randomized trials: an extension of the CONSORT statement. *Ann Intern Med* 2004; 141(10):781-788.
