## Supplementary material for "Triaging and Referring In Adjacent General and Emergency Departments (the TRIAGE trial): a cluster randomised controlled trial": Statistical Analysis Plan

**S14 Statistical Analysis Plan.**

| 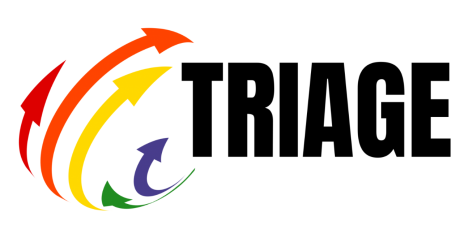 | 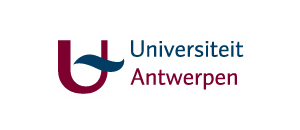 |
| --- | --- |

Statistical analysis plan for the TRIAGE-trial: Triaging and Referring In Adjacent General and Emergency departments (the TRIAGE-trial), a cluster randomised controlled trial.

Version: 1.0

Date: April 2021

Based on the study protocol version 2.1 December 2018

Statistician: Jarl K. Kampen

Principal Investigators: H. Philips, V.Verhoeven, and S. Morreel

1. Content Table

### 1. Introduction

**The problem: I am feeling ill during the weekend, where should I go?**

When confronted with an unexpected illness during the weekend, patients can go either to the emergency department (ED) of a hospital or to the general practitioner (GP) on call. In Flanders, the GPs often organise this on call service for a specified region in a central location called General Practice Cooperatives (GPC, “Wachtpost” in Dutch). Patients do not know the characteristics of these different out of hours (OOH) services and find it hard to estimate the urgency of their own complaints. Therefore, many patients with presentations suitable for primary care go directly to the ED. This leads to additional costs for both government and patient, long waiting times at the ED and a high workload for the emergency physicians in Flanders.

**Aim of this project: we will advise you where to go**

The aim of this project is to deliver the most appropriate care for patients presenting at the ED. To achieve this objective a triage nurse will assess all patients presenting at an ED and advise them on the most appropriate point of care: GPC or ED. Triage is ‘the sorting out and classification of patients to determine priority of need’. Extended triage is triage combined with allocation to either ED or GPC, which is new in Flanders.

Many EDs in Flanders use the Manchester Triage system (MTS) in their regular care for triage within the ED, to sort patients according to the urgency of their presentation. We developed an extended MTS (eMTS) which foresees allocation to either ED or GPC. Scientific research is crucial to evaluate such triage systems before implementation is possible. Our study is a cluster-randomised trial. During one year, we will perform extended triage using the eMTS at one ED/GPC collaboration. Every four weekends, one weekend will serve as a control: the patient does not get an allocation advice.

This project is a realisation of a multi-disciplinary university research consortium and a collaboration of an ED and GPC as the research site.

At the end of this trial, we will deliver the first validated tool to perform extended triage and referral to primary care during OOH. Afterwards we aim to implement this tool throughout Flanders. Ultimately, we want to achieve a more efficient delivery of OOH care for Flemish patients with a cost reduction for the Flemish healthcare system.

**Scientific methodology of this project: is this service at this time the most appropriate for me?**

Three aspects determine the most appropriate OOH service for a specific patient at a specific time:

- Medical aspects: is this service capable of handling the medical problem in an efficient and qualitative way? Is it safe?
- Financial aspects: is it the most cost effective service for both government and patient?
- Process aspects: can the delivered service satisfy the perceived needs of patients and staff? What are the major barriers and facilitators? What can we learn from reported incidents? How can we improve safety? This aspect is only briefly described in this protocol, as we will make a separate protocol.

For more profound information on the trial we refer to the research protocol of the medical aspect.

### 2. Study objectives

The objective of the current study was to determine the efficiency and safety of a nurse-led triage system streaming low risk patients who present at an ED to the adjacent GPC. See protocol for more details.

### 3. Study design

#### 3.1 Overview

Single centre, cluster randomized trial with weekends and bank holidays serving as clusters.

#### 3.2 Study population

We systematically assessed all patients presenting at the ED during the weekend for inclusion. By presentation at the ED we mean the patient is registered by the receptionist of the ED as a patient. Patients who just pass the ED to enter the hospital or who go directly to a medical department (e.g., obstetrics) are thus not considered for inclusion. See protocol for details.

#### 3.3 Inclusion criteria

The availability of a Belgian citizen national insurance number is the only inclusion criterion.

#### 3.4 Exclusion criteria

Patients arriving at the ED by an ambulance with a doctor or nurse (in Dutch called MUG or PIT) were excluded (these patients have already passed through a sort of triage). Pleas notice there was a flaw in the study protocol: patients referred by a health care professional were not explicitly mentioned as an exclusion but are always triaged to the ED and as such they are excluded as well. We have corrected this in the study paper and reported in the “changes to the protocol” section of the supplementary material.

#### 3.5  Sample size

If we only look at the primary objective, straightforward power computations will tell that two weekends (one with, and one without intervention) are sufficient to provide empirical evidence of a statistically significant shift. However, as we want to know the determinants of this proportion and perform safety monitoring (including incident analysis), we need observations over a longer period of time. We do not know the impact size of the determinants that might act as confounders in this context. Because the eMTS is risk adverse, safety issues will be rare and a long study period is required to identify and study possible safety issues. Finally, we expect nurses will need to build up experience with the eMTS before optimal results can be expected. For these reasons, we have chosen to include a large convenience sample of ten months.

#### 3.6  Randomization

One out of every four weekends served as a control. We will assign the control weekends using randomisation software, taking into account bank and school holidays. We were not be able to blind the participants, the staff members nor the researchers. The head nurse and one assistant were aware of the randomisation and communicated the allocation of the upcoming weekends to the ED staff a few hours before its start. The GPC staff was not informed about the randomisation but could find it out during their shift through patient contacts. Patients were not blinded for the intervention.

#### 3.7 Study schedule

See protocol

#### 3.8  Study duration

Start: 4/01/2019 (test phase, the trial starts on 01/03/2019

End: 31/12/2019

Final end of study: 31/12/2019

Follow-up period: total stay at ED or GPC

Dropout: only when a patients leaves the ED or GPC after triage he or she is considered drop-out, the expected rate is low. These participants will be called left without being seen.

### 4. Study endpoints

#### 4.1 Primary endpoint

Difference in Proportion of study patients seen by the GP between intervention and control weekends.

#### 4.2 Secondary endpoint

- Proportion of patients who are not compliant to a correct GPC advice

#### 4.3 Other endpoints

- Determinants of the primary outcome
- Referral rate back for patients allocated to the GPC
- Admissions to the study hospital
- Performance of the eMTS as an instrument to detect patients suitable for primary care
  - Sensitivity and specificity were not reported (see changes to the study protocol)
  - Positive and negative likelihood ratio for the detection of patients suitable for primary care by a triage nurse

### 5. Sequence of planned analyses

#### 5.1 Interim analyses

SAEs are monitored on a regular basis but are not measurable by our endpoint (hence patients with severe diseases are admitted to the ED all the time, the influence of the intervention will generally be very unlikely). These SAE’s form part of a process analysis described elsewhere. All participating health care workers and the hospital’s ombudsperson were asked to report all (near) incidents possibly related to the study to the same working group.

One and six months after the start of the trial, the research team presented interim results to the working group that prepared the study. After the second meeting, the nurses were asked to provide a motivation in free text whenever they chose the discriminator “GP Risk” (a pre-specified risk or a personal opinion).

#### 5.2 Final analyses and reporting

All final planned analyses are conducted only after the end of the trial.

### 6. Statistical methods

#### 6.1  Analysis principles

Patients who withdraw consent for use of their data will not be included in any analysis.

Two-sided 5% significance levels will be used to identify statistically significant results. All confidence intervals reported will be 95% confidence intervals.

#### 6.2  Incomplete follow-up, missing data and outliers

Given the nature of the data it was not possible to make a detailed plan about these subjects in advance. As an alternative all data cleaning is reported here. Ultimately, 921/9964 records were cleaned for at least one variable. All corrected variables were stored in a separate table. The final study database was constructed on the basis of the raw data overruled by this separate table when necessary.

#### 6.3 Missing outcome data

- There was no withdrawal of consent
- The primary outcome was ultimately known for most patients except when the secondary outcome (triage advice) was also missing

#### 6.4 Missing baseline covariates

These have been reported in table 1 of the study paper.

#### 6.5 Outliers

No variables with relevant outliers are reported. Notice that most variables are categorical

#### 6.6 Data cleaning

Given the nature of the collected data (routine medical records), it was not possible to make a data cleaning plan in advance. Hereafter we report it as it was carried out.

##### 6.6.1 Primary outcome

Because the nurses did not always fill in all variables concerning triage (advice GPC or ED and agreement yes or no), data cleaning was necessary. These data were more often missing when the patient was ultimately seen at the GPC compared to the ED. All patients with a GPC contact within a time frame of eight hours before and after the start of the triage were included in this data cleaning. All these records were analyzed by and automated decision rule and manually by the first author. The result was double checked by either author VV or HP. The routine medical data contain variables not studied in the current study but that do give information about the primary outcome. For example, there is a variable “destination” in which the nurse should write the destination of the patient after the ED (ward, intensive care ,… but also GPC) and at the GPC there is a variable “origin” (home, ED or telephone triage). Additionally, the time of presentation at the ED and GPC and the variable “destination” at the GPC also give valuable information. Records which remained unclassifiable after this data cleaning were examined by the head nurse of the study which was able to categorize all of them.

##### 6.6.2 Patients within the primary outcome referred back to the ED

These patients were identified using the data from the GPC (variable “destination”) but checked manually as well as described in 6.2.4.2. In order not to include them twice, when appropriate, the second triage (after GPC consultation) was removed. Sometimes the nurses did not write down a second advice but changed the first one, in that case it was restored back to the original one.

##### 6.6.3 The variable “Accepts allocation advice” and the variable “Allocation advice”

Because the nurses did not always fill in all variables concerning triage, data cleaning was necessary. The study database contains a free text variable which was read by the first author for all cases. When this variable was not in line with the allocation advice or its acceptance, this variable was overruled. A common example: the nurse registered a patient with the advice “GPC” and the acceptance “No” but wrote down a motivation why the nurse overruled the advice: “I felt this patient was in need of urgent imaging” 🡺 this case was overruled to “ED” and “Yes”.

##### 6.6.4 Variable allocation advice is missing but the discriminator “GP risk was chosen”

As choosing the discriminator GP risk is another way to register an ED advice, these records were completed with an ED allocation advice.

##### 6.6.5 Under registration of arrival by ambulance with physician or nurse

During data cleaning by the first author, all free text fields were read. Occasionally this contained information like “transferred to the ED by ambulance equipped with physician” but this information was not registered correctly. These false registration were overruled

##### 6.6.6 Patients with a high urgency triage but allocated to the GPC

Rarely a patient within the urgency categories one to three was registered with a GPC advice. After thoroughly examining all these records by the first author, some of them were judged as a registration error as all contained a report from the ED and none contained any other information making a true GPC allocation likely.

##### 6.6.7 Patients with a high urgency triage but no allocation advice available

Se 6.2.4.6: as these patients were extremely unlikely to get a GPC advice, this missing advice was overruled to “ED”.

##### 6.6.8 Empty records

Entirely empty records were removed from the study database, records containing many missing values were checked by the head nurse, most of the turned out to be double registrations or patients who left even before triage.

##### 6.6.9 Patients registered as refusing a GPC advice but seen by at the GPC and not seen at the ED

These records were checked manually by the first author and the advice was overruled when all other variables and the free text fields clearly indicated a GPC advice.

##### 6.6.10 Patients refusing an ED advice

As it seems very unnatural to present oneself at the ED and afterwards refusing the advice to stay at the ED, all of these records were controlled manually and transferred to the head nurse when unclear. Only fourteen real refusals of an ED advice remained. As these patients were not the focus of the study and not all of them actually went to the GPC, they were considered as left without being seen.

##### 6.6.11 Patients apparently triaged by a nurse that was not allowed to triage

Only nurses with at least one year of experience at the study hospital were allowed to triage. Even though the researchers found some cases of patients triaged by nurses which were not allowed to do so. After consulting the head nurse, it turned out that these nurses never triage but sometimes write down the result of their colleague’s triage or forgot to log out of a computer. For these cases, the nurse ID was overruled to “missing” as we do not know who was the true triage nurse.

##### 6.6.12 Patients triaged by a physician and not a nurse

In the variable Nurse ID, sometimes the ID of a physician appeared. After consulting the head nurse, it turned out that some physicians occasionally see a patient before triage and handle these patients alone. Because these patients are not in the scope of this trial (which is nurse led triage), they were removed from the study database (they were not even assessed for eligibility and thus do not appear in the report).

##### 6.6.13 Hospitalizations

For the first six months of the study, we already had the financial data. As this data contains a variable hospitalized (yes or no), this data was used to improve this variable in the medical study data.

#### 6.7 Data transformations

Data transformations were not necessary.

#### 6.8  Multiple comparisons and multiplicity

In the CHAID analysis, Bonferroni-Holms correction was used.

#### 6.9  Data management and analysis software

Data have been collected, cleaned, and managed in JMP® version 15. Alle analysis have been performed in JMP® except the CHAID analysis which has been executed in SPSS version 25 and the regression model which was built in Jamovi version 1.6 with the module GAMLj: General Analyses for the Linear Model in Jamovi.[1] The epiR package in R version 4.0 was used to calculate positive and negative predictive values with a 95% CI. [2]

All scripts used are available upon reasonable request as are the outputs from the software.

### 7. Adherence and protocol deviations

True dropouts from the intervention did not occur as none of the patients ended their triage consultation prematurely. Drop-out after the intervention have been reported as left without being seen, a more common and precise term in the field of research about emergency care.

There were no protocol deviations, all patients/weekends received the allocated intervention. During one holiday (15/08/2019), the study was not started so no data is available about this day. This holiday was originally randomised to the intervention group. These patients (n= unknown) did not receive the studied intervention, neither the control intervention so they were not studied and consequently, the entire report remains an intention to treat analysis.

### 8. Trial population

The eligibility population and all relevant patient groups have been reported as a CONSORT-flow chart. Only patients triaged by a nurse were eligible for inclusion so the rare patients directly triaged by a physician were not incorporated (see data cleaning for more details).

### 9. Statistical analyses

#### 9.1  Availability of statistical outputs and scripts

The scripts used for the data management, data cleaning and data analysis are available upon reasonable request as are outputs from the statistical software.

#### 9.2  Patient characteristics and baseline comparisons

Demographic and other baseline characteristics have been summarized by treatment group. For categorical variables, frequencies and percentages have be reported. Missing values have been reported. Continuous variables have been summarized as mean with standard deviation.

Comparisons of demographic and baseline characteristics between the treatment groups have be conducted to assess the degree to which comparability of the groups was achieved by the randomization. For categorical variables chi-squared test or Fisher exact test (when numbers are low) have been used. For continuous variables, a t-test was used as appropriate.

#### 9.3  Analysis of the determinants of the primary outcomes

The difference between control and intervention weekends with respect to the primary outcome were reported in terms of the odds ratio and confidence interval. Logistic regression was used to calculate odds ratios of the dichotomous outcomes across multi-level categorical variables. The category with a mean primary outcome closest to the overall primary outcome (9.5%) was set as the reference category. Those variables found significant (two tailed alpha = 0.05) in the univariate analysis or believed to be of specific interest or prone to major interactions were incorporated in the multivariate analysis. This multivariate analysis (both for the primary and the secondary outcome) was started by creating three CHAID decision trees. [3, 4] Decision tree methodology is a useful data mining method for developing prediction algorithms for a target variable. This method classifies a population into branch-like segments that construct an inverted tree with a root node, internal nodes, and leaf nodes. The algorithm is non-parametric and can efficiently deal with large, complicated datasets and can accept missing values.[5] Decision trees based on Bonferroni-Holms corrected chi-squared test were constructed separately for study tool (urgency category, presentational flowchart category), patient characteristics (type of admission, sex, age, living nearby, social status) and time of presentation (weekend, time period, subjective crowding). The analysis was validated using the K-fold cross validation method. The model was built with the following specifications: a maximum tree depth of four, minimum cases in parent nodes of 50, minimum of cases in child notes of 25, missing values analysed as a separate category, and an alpha-value for splits at 0.05. Because the proportion of the primary outcome was below 50% in all branches for the primary outcome and in the majority of branches for the secondary outcome, the misclassification risk has not been reported as it was by definition equal or very close to the outcome studied.

A final model was fitted using the significant nodes as they turned out in the separate analyses. To compare the primary outcome during the study period to the year 2020, an unpaired samples student’s t-test was used.

#### 9.4  Analysis of secondary endpoints

The same as the primary endpoint but with an additional variable: intervention (intervention or control). No regression model was constructed as it would complicate the paper without adding clinically relevant information.

#### 9.5  Analysis of safety endpoints

The was no formal analysis of adverse events but as a surrogate marker of safety, the diagnostic properties of the studied tool as an instrument to detect primary care patients, the admissions to the study hospital and the referral rate back to the ED have been assessed.

#### 9.6 Diagnostic properties of the studied tool

Because the numbers of patients who refused the allocation advice was rather low, these patients were excluded from this analysis as well as the patients without a known allocation.

During intervention weekends: for patients within the primary outcome (n=599), the gold standard is the advice of the GP. Patients referred back to the ED were considered “false positive”, the others true positives. Positive and negative predictive values were calculated in R using the epiR package). For patients with an accepted advice ED (n=5452), the gold standard is the opinion of the ED physician: false negative when these patients were suitable for primary care or true negative when these patients were not. As the missing values for this gold standard were unequally distributed (by definition never missing at the GPC, often missing at the ED), it was not possible to calculate meaningful and reliable sensitivity and specificity values:

| Triage: Suitable for primary care | Golden standard: Yes | Golden standard: no | Missing |
| --- | --- | --- | --- |
| Yes | 575 | 24 | 0 |
| No | 797 | 1196 | 2000 |

During control weekends it is possible to make this calculation as there is only one gold standard but the authors choose not to report this as it would only confuse the readers without adding clinically relevant information.

#### 9.7 Referrals back to the ED and admissions to the study hospital

These dichotomous variables were analysed exactly like the primary and secondary outcome

#### 9.8 Exploration of the primary outcome after the trial

The triage process continued after the trial, but the ED stopped registration of some variables while others were registered less strictly making extraction of the data from the ED unreliable. Additionally, it was not feasible to continue the data cleaning efforts after the trial and the total ED population (inclusions in the study) was not available making calculation of the proportion of the primary outcome impossible.

To make an exploration of the primary outcome after the study period, the number of patients allocated to the GPC as noted by the receptionists of the GPC was extracted directly from the GPC’s software (Mediris®2.4) both for the study period and one year afterwards by the first author. Because the COVID-19 pandemic disrupted the Belgian healthcare system (leading to a large decline of the demand for acute care unrelated to COVID-19) during two waves, the months April, November, and December 2020 were excluded. [6]

Because this is another way to measure the same primary outcome, this methodology was also applied to the study period itself (intervention group only). An unpaired t-test was used to compare both groups.

### 10. References

1.	The Jamovi Project. Jamovi. 1.6 ed2021.

2.	R development Core Team. R: A language and environnement for statistical computing. 4.0 ed: R Foundation for Statistical Computing; 2021.

3.	Kass GV. An Exploratory Technique for Investigating Large Quantities of Categorical Data. Journal of the Royal Statistical Society: Series C (Applied Statistics). 1980;29(2):119-27.

4.	Lemon SC, Roy J, Clark MA, Friedmann PD, Rakowski W. Classification and regression tree analysis in public health: methodological review and comparison with logistic regression. Annals of behavioral medicine : a publication of the Society of Behavioral Medicine. 2003;26(3):172-81.

5.	Song Y-Y, Lu Y. Decision tree methods: applications for classification and prediction. Shanghai Arch Psychiatry. 2015;27(2):130-5.

6.	Morreel S, Philips H, Verhoeven V. Organisation and characteristics of out-of-hours primary care during a COVID-19 outbreak: A real-time observational study. PLoS One. 2020;15(8):e0237629.
