## Supplementary Material for "Triaging and Referring In Adjacent General and Emergency Departments (the TRIAGE trial): a cluster randomised controlled trial"

### Supplementary table 1

Bivariate analysis of the secondary outcome (all participants excluding those with a missing triage advice). For categorical variables with more than four categories, the categories with the highest and lowest primary outcome are reported.

| **Determinant** | **N** | | **Mean secondary outcome** | **DF** | **Category** | **Estimate** | **Wald Chi²** | **P-value** | | **Odds ratio (95%CI)** |
| --- | --- | --- | --- | --- | --- | --- | --- | --- | --- | --- |
| **Patient’s presentation** | | | | | | | | | | |
| MTS urgency category* | 4823 | | 26.1% | 1 | 4: Standard |  | | | | 1 |
|  |  |  |  |  | 5: Non-urgent | 1.65 | 50.8 | <0.01 | | 5.18 (3.30 to 8.15) |
| MTS flow chart category | 7978 | | 15.7% | 14 | Unwell adult |  | | | | 1 |
|  |  |  |  |  | ORL Complaints | 1.13 | 64.2 | <0.01 | | 3.08 (2.34 to 4.06) |
|  |  |  |  |  | Chest pain | -2.84 | 23.3 | <0.01 | | 0.06 (0.02 to 0.19) |
| **Patient characteristics** | | | | | | | | | | |
| Age | | 8038 | 15.8% | 5 | 0-7 years | 0.21 | 3.7 | **0.05** | | 1.23 (1.00 to 1.52) |
|  |  |  |  |  | 8-24 years | 0.25 | 6.6 | 0.01 | | 1.28 (1.06 to 1.55) |
|  |  |  |  |  | 25-39 years | 0.19 | **4.0** | 0.05 | | 1.21 (1.00 to 1.45) |
|  |  |  |  |  | 40-54 |  | | | | 1 |
|  |  |  |  |  | 55-74 | -0.39 | 11.1 | <0.01 | | 0.68 (0.54 to 0.85) |
|  |  |  |  |  | >74 | -1.02 | 41.6 | <0.01 | | 0.36 (0.27 to 0.49) |
| Admission type | 8034 | | 15.8% | 1 | Walk-in |  | | | | 1 |
|  |  |  |  |  | Arrived by ambulance | -1.71 | 131.5 | | <0.01 | 0.18 (0.14 to 0.24) |
| Sex | 8038 | | 15.8% | 1 | Female |  | | | | 1 |
|  |  |  |  |  | Male | -0.08 | 1.61 | 0.20 | | 0.93 (0.82 to 1.04) |
| Residence | 8012 | | 15.8% | 1 | Nearby |  |  | | | 1 |
|  |  |  |  |  | Not living nearby | -0.28 | 16.8 | <0.01 | | 0.75 (0.65 to 0.86) |
| Socioeconomic status | 6788 | | 16.7% | 1 | Normal |  | | | | 1 |
|  |  |  |  |  | Low | 0.45 | 45.1 | <0.01 | | 1.57 (1.38 to 1.79) |
| **Timing of presentation** | | | | | | | | | | |
| Intervention | 8038 | | 15.8% | 1 | Intervention |  | | | | 1 |
|  |  |  |  |  | Control | 0.71 | 99.4 | <0.01 | | 2.03 (1.77 to 2.34) |
| Weekend | 8038 | | 15.8% | 46 | 16/08/2019-19/08/2019 |  | | | | 1 |
|  |  |  |  |  | 26/04/2019-29/04/2019 | 1.02 | 13.0 | <0.01 | | 2.78 (1.59 to 4.84) |
|  |  |  |  |  | 13/12/2019-15/12/2019 | -1.20 | 9.0 | <0.01 | | 0.30 (0.14 to 0.66) |
| Time period | 8038 | | 15.8% | 2 | Day |  | | | | 1 |
|  |  |  |  |  | Evening | -0.17 | 5.5 | 0.02 | | 0.84 (0.73 to 0.97) |
|  |  |  |  |  | Night | 0.08 | 1.01 | 0.31 | | 1.09 (0.93 to 1.27) |
| Subjective crowding at the ED | 3279 | | 13.3% | 2 | Normal |  | | | | 1 |
|  |  |  |  |  | Quiet | 0.60 | 15.9 | <0.01 | | 1.82 (1.36 to 2.44) |
|  |  |  |  |  | Busy | 0.28 | 3.7 | 0.06 | | 1.32 (2.99 to 1.76) |
| **Nurse characteristics** | | | | | | | | | | |
| Nurse | 7635 | | 16.4% | 21 | Nurse 9 |  | | | | 1 |
|  |  |  |  |  | Nurse 7 | 0.89 | 24.3 | <0.01 | | 2.46 (1.72 to 3.52) |
|  |  |  |  |  | Nurse 19 | -0.73 | 7.4 | 0.01 | | 0.48 (0.28 to 0.81) |

DF: degrees of freedom
MTS: Manchester Triage System
ED: Emergency Department
ORL: Otorhinolaryngology
*: Only for urgency categories four and five because the primary outcome was zero in the other categories

### Supplementary table 2

Characteristics of patients referred back to the ED after triage to the GPC

| Age  Range | Sex | eMTS presentational flow chart | GP diagnosis | Hospital care | Admission |
| --- | --- | --- | --- | --- | --- |
| 10-20 | MALE | Falls | Concussion | Imaging | No |
| 10-20 | FEMALE | Back pain | Sciatica | None | No |
| 30-40 | FEMALE | Abdominal pain in adults | Localised abdominal pain | None | No |
| 10-20 | MALE | Limb problems | Musculoskeletal injury | None | No |
| 10-20 | FEMALE | Unwell adult | Unwell | Imaging | No |
| 0-10 | MALE | Headache | Headache | Imaging | No |
| 10-20 | FEMALE | Abdominal pain in adults | In labour | None | No |
| 80-90 | MALE | Wounds | Cut/laceration | Imaging | No |
| 40-50 | FEMALE | Unwell adult | Headache | Monitoring of vital signs | Yes |
| 70-80 | MALE | Abdominal pain in adults | Kidney stones | None | No |
| 30-40 | FEMALE | Abdominal pain in adults | Unspecified illness | Monitoring of vital signs | No |
| 10-20 | MALE | Headache | Symptoms of the nervous system | None | Yes |
| 10-20 | FEMALE | Abdominal pain in adults | Kidney stones | None | No |
| 20-30 | FEMALE | Limb problems | Musculoskeletal injury | None | No |
| 40-50 | FEMALE | Limb problems | Unspecified fracture | None | No |
| 20-30 | FEMALE | Headache | Headache | None | No |
| 20-30 | FEMALE | Limb problems | Musculoskeletal injury | None | No |
| 20-30 | FEMALE | Abdominal pain in adults | Generalised abdominal pain | None | No |
| 0-10 | MALE | Unwell baby | Fever | None | No |
| 0-10 | MALE | Abdominal pain in children | Unspecified illness | None | Yes |
| 50-60 | FEMALE | Headache | Unspecified illness | None | No |
| 30-20 | MALE | Abdominal pain in adults | Unspecified illness | None | No |
| 0-10 | FEMALE | Unwell child | Fever | Monitoring of vital signs | No |
| 20-30 | FEMALE | Back pain | Symptoms/complaints regarding back pain | None | No |

MTS: Manchester Triage System
GP: General Practitioner

### Supplementary table 3

Generalised Mixed Model for the primary outcome

| Determinant | Chi square | Df | P-value |
| --- | --- | --- | --- |
| MTS flow chart category | 567.3 | 15 | <0.01 |
| Admission type | 51.2 | 1 | <0.01 |
| Nurse identifier | 144.5 | 22 | <.0.01 |
| Subjective crowding | 15.4 | 3 | <0.01 |
| Residence | 11.5 | 2 | <0.01 |
| Socioeconomic status | 21.9 | 2 | <0.01 |
| Time period | 42.5 | 3 | <0.01 |
| Age | 12.2 | 5 | 0.032 |

MTS: Manchester Triage System

Df: degrees of freedom

### Supplementary figure 1: Primary Outcome - Chi-square automatic interaction detection (CHAID) decision tree of the study tool parameters

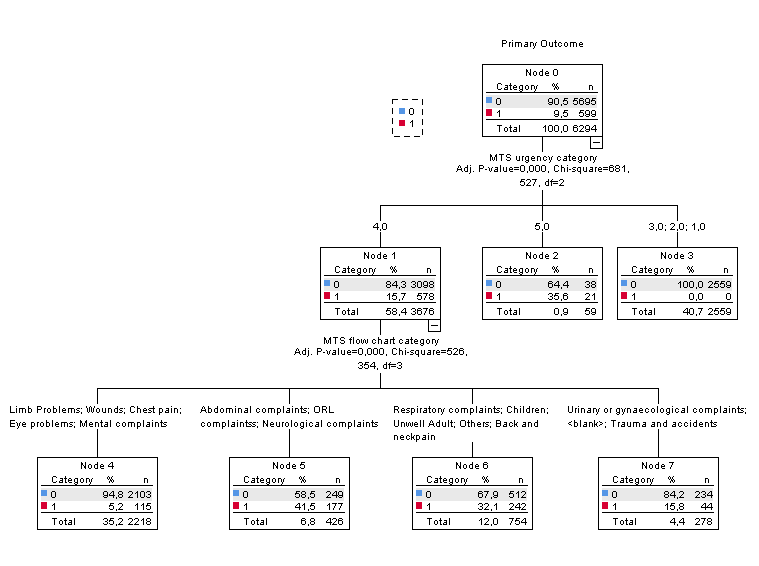

ED: Emergency Department
GPC: General Practice Cooperative
MTS: Manchester Triage System
ORL: Otorhinolaryngology
<blank>: no MTS flow chart was registered

### Supplementary figure 2: Primary Outcome - Chi-square automatic interaction detection (CHAID) decision tree of the patient characteristics

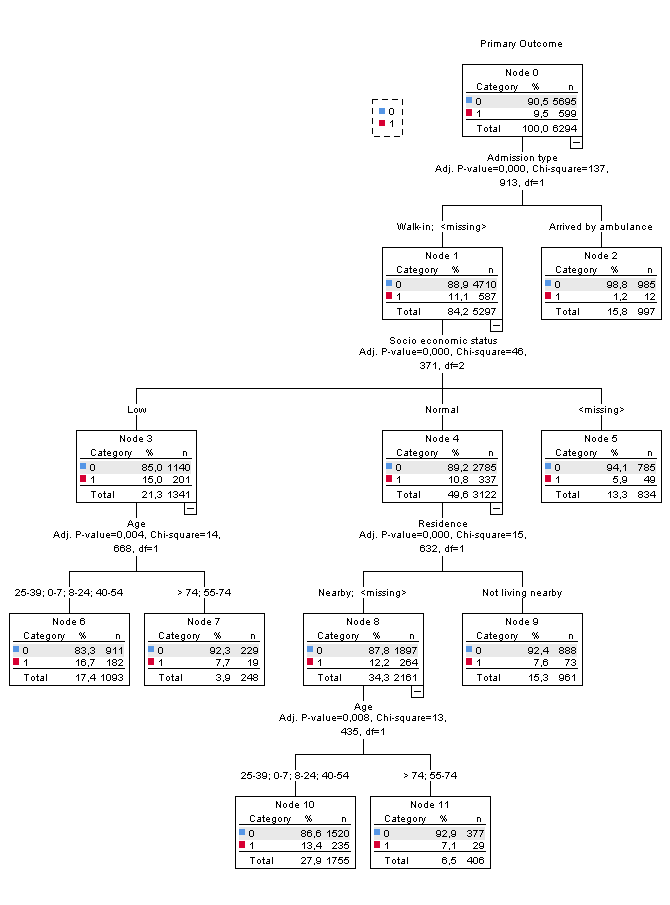

### Supplementary figure 3: Primary Outcome - Chi-square automatic interaction detection (CHAID) decision tree of the timing of presentation

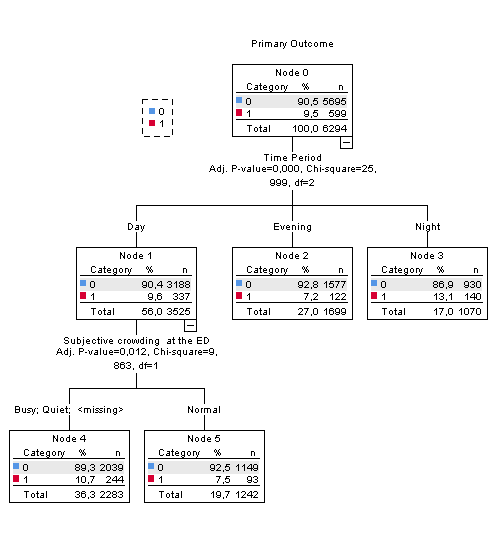

### Supplementary figure 4: Secondary Outcome - Chi-square automatic interaction detection (CHAID) decision tree of the study tool parameters

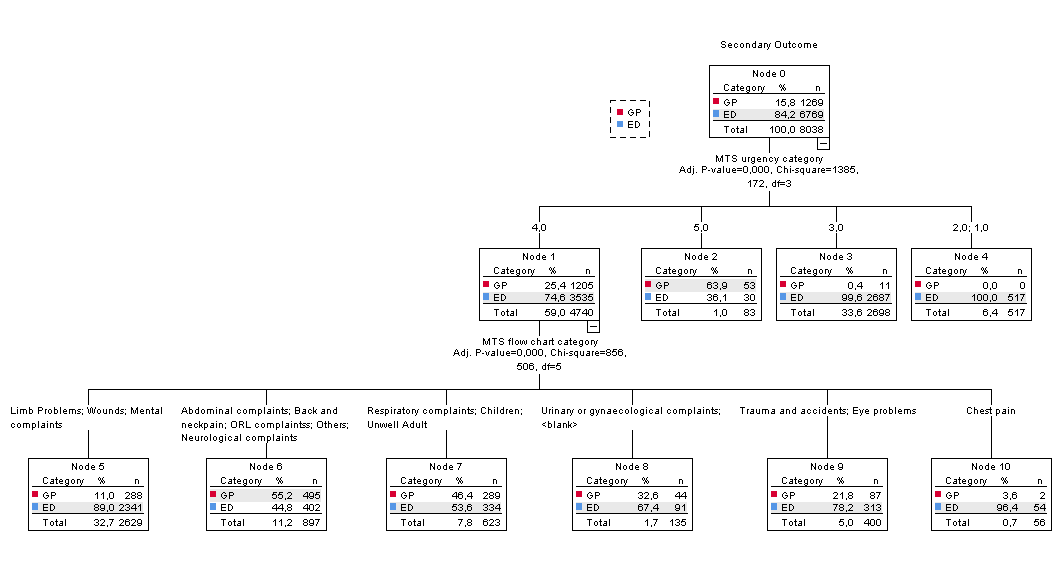

### Supplementary figure 5: Secondary Outcome - Chi-square automatic interaction detection (CHAID) decision tree of the patient characteristics

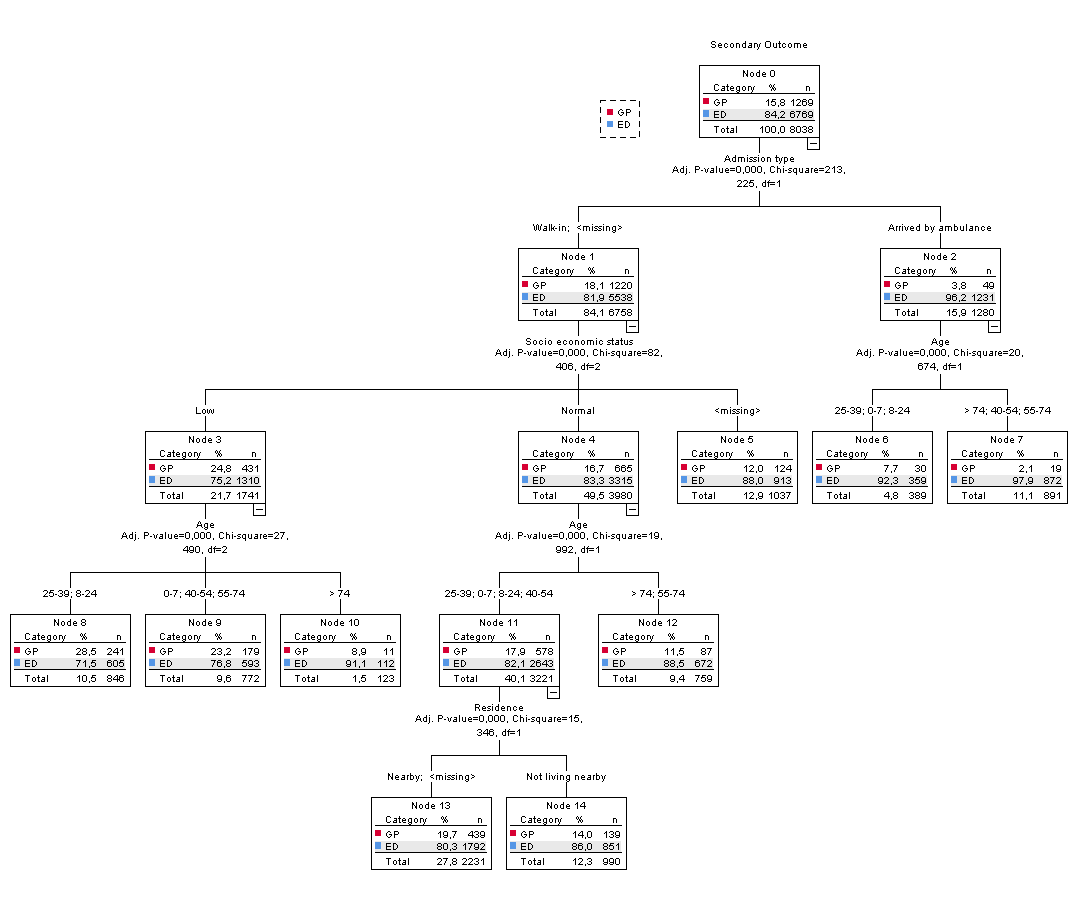

### Supplementary figure 6: Secondary Outcome - Chi-square automatic interaction detection (CHAID) decision tree of the timing of presentation

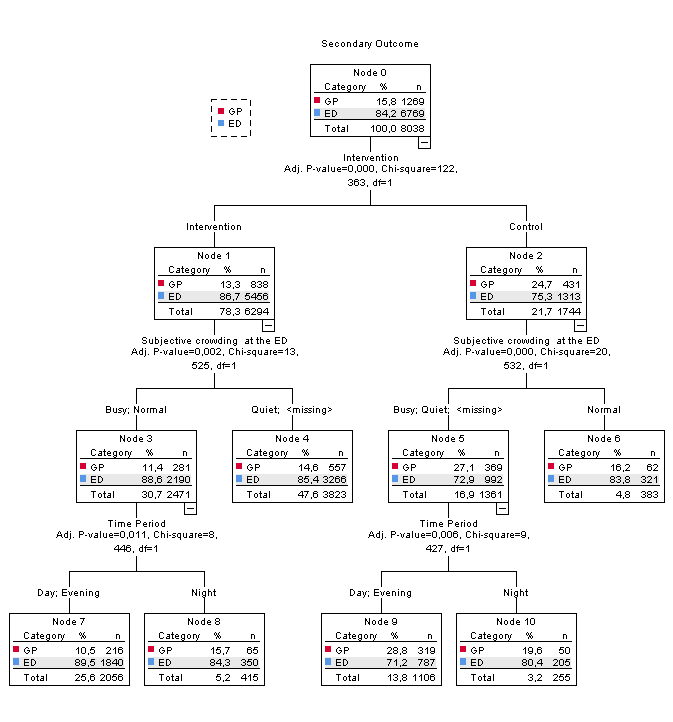

Supplementary figure 7: Secondary Outcome - Chi-square automatic interaction detection (CHAID) combined decision tree
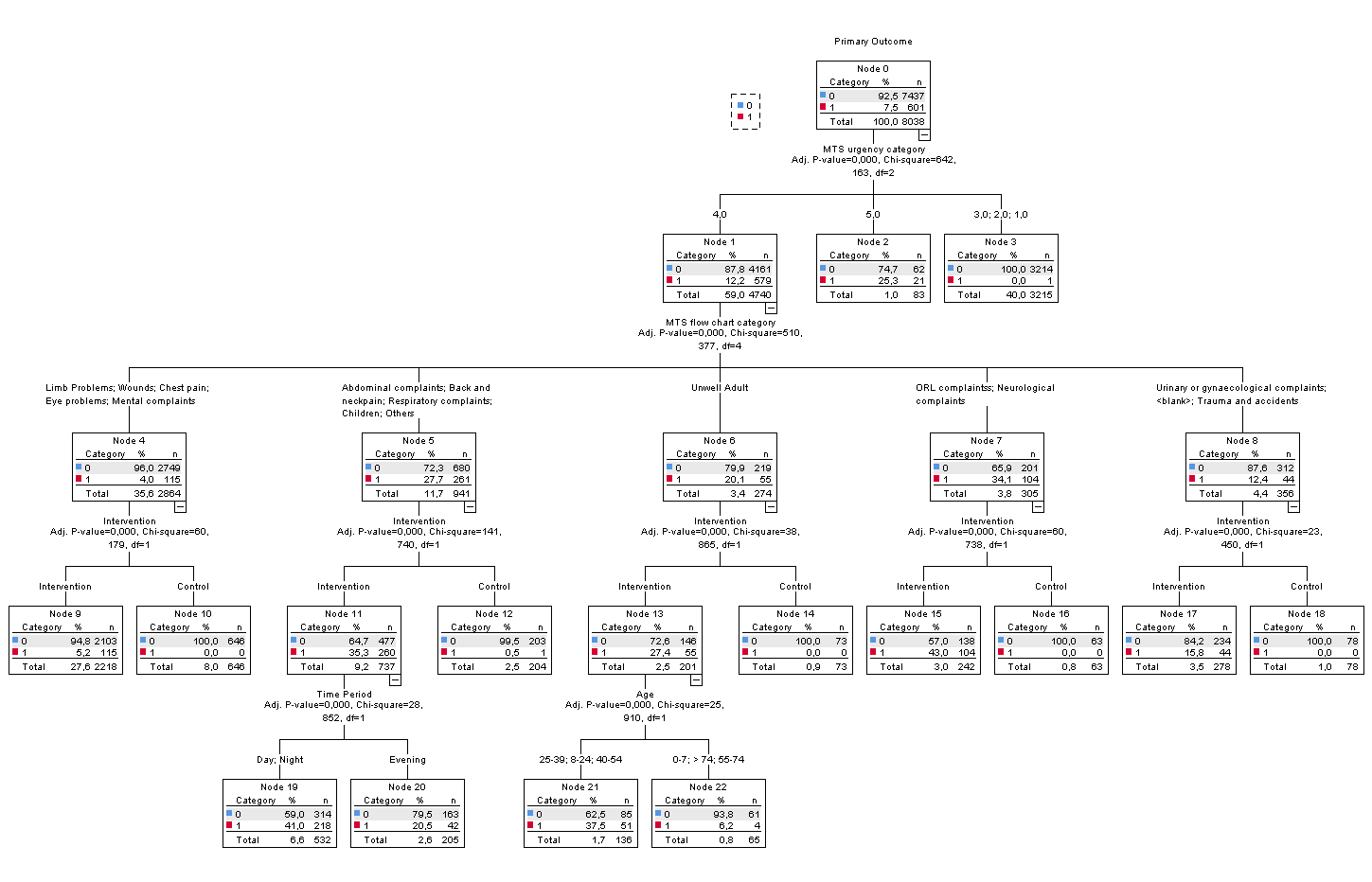

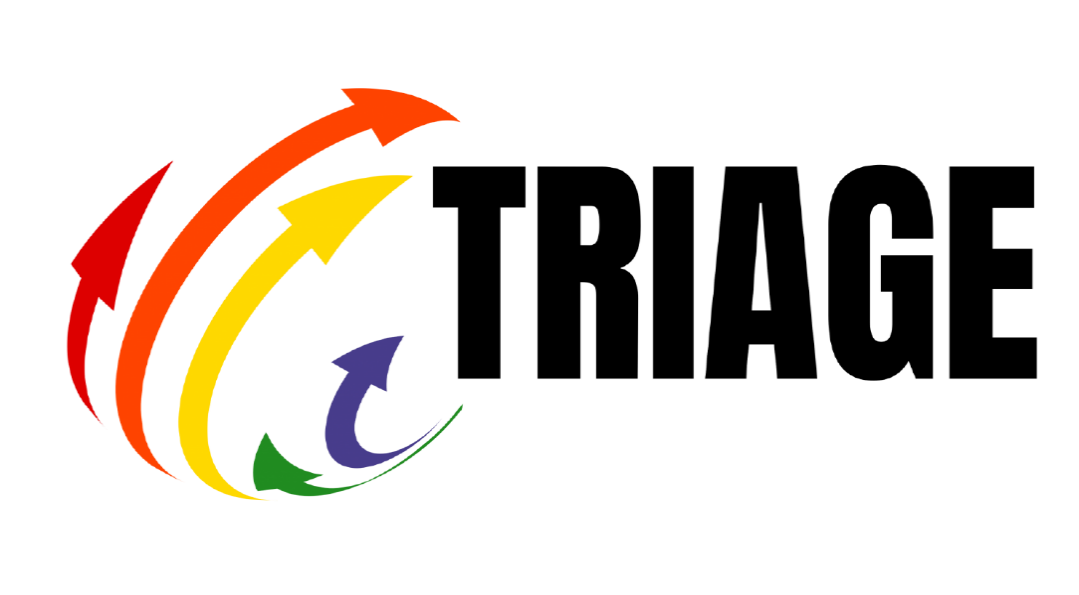

### Research protocol

Triaging and Referring In Adjacent General and Emergency departments (the TRIAGE-trial): a cluster randomised controlled trial

Version: 2.1 December 2018

##### Administrative information

**Full title:** Triaging and Referring In Adjacent General and Emergency departments (the TRIAGE-trial): a cluster randomised controlled trial

**Short Title:** the TRIAGE-trial: Triaging and Referring In Adjacent General and Emergency departments

**Registration:** we will register the trial on clinicaltrials.gov see also Attachment 1: World Health Organization Trial Registration Data Set

**Protocol version:** 1.0 August 2019

**Financial support:** this trial is sponsored by the Research Foundation - Flanders (FWO). Grant Number T000718N. The sponsor is not involved in the study-design, data collection, management, analysis and interpretation, writing of the report nor publication of the results.

**Authors:**

The TRIAGE-trial is a complex healthcare intervention with involvement of many stakeholders:

- The Centre of General Practice of the University of Antwerp (part of research group ELIZA), coordination centre
  - Stefan Morreel^1,2^: main researcher, principal author of this entire document unless stated otherwise
  - Hilde Philips^1,2^: promotor of the entire trial, data protection officer
  - Veronique Verhoeven^1,2^: promotor of the entire trial, spokesperson
- The Emergency Department of the Antwerp University Hospital (part of research group ASTARC)
  - Koen Monsieurs^1^: promotor of the medical aspect
  - Joo-Ree Melis^2^: study-nurse
- Department of General Economics of the University of Antwerp
  - Diana De Graeve^1^: promotor for the financial aspect
- Local clinical committee (“werkgroep triage”):
  - Sander Naeyaert^2^: chief nurse of the Emergency Department (ED) of the general hospital AZ Monica Deurne. Several emergency nurses have helped him for the TRIAGE-trial. He receives support from the FWO in the form of an extra triage nurse during the trial (Ragna Verlent).
  - Arnoud Bonemeyer and Mark Timmermans: emergency physicians at the ED of AZ Monica Deurne
  - Edwin Vanbeveren: head of the General Practice Cooperative (GPC) “Antwerpen Oost”
  - Guido Michielsen and Lotte Fivez: Board members of the GPC “Antwerpen Oost”

^1^: co-author of this document

^2^: receives (indirect) FWO support

##### Introduction

###### Dutch Abstract

Available upon request

###### English abstract

**Introduction**: Patients who might also go to the general practitioner (GP) frequently consult emergency departments (ED). This leads to additional costs for both government and patient and a high workload for emergency physicians in Flanders. The Belgian government wants to address this problem by improved collaboration between EDs and general practice cooperatives (GPCs).

**Intervention**: Patients presenting at the ED during out-of-hours (OOH) will be triaged and allocated to the most appropriate service. For this purpose the Manchester Triage System (MTS) which is commonly used in Flemish hospitals, will be extended (eMTS). By doing so a trained nurse will be able to diverge suitable patients towards the GPC.

**Methodology**: We will conduct a cluster RCT in which eligible ED patients will be diverged to the GPC using the eMTS. We will collect data using our operational anonymous database for OOH care (iCAREdata). We will study the use of the eMTS, the effectiveness and effects of triage, work load changes, epidemiology at both departments, patient safety, health insurance (HIS) and patient expenditures. Furthermore, facilitators and barriers will be studied and an incident analysis of problem cases will be performed.

**Outcome**: The primary outcome is the proportion of patients who enter the ED and are handled by the GP after triage. Secondary outcome measurements are related to safety: referral rate to the ED by the GP, proportion of patients visiting the ED again within two weeks, proportion of patients not following the triage advice and file review for selected patients.

###### Overview

**The problem: I am feeling ill during the weekend, where should I go?**

When confronted with an unexpected illness during the weekend, patients can go either to the emergency department (ED) of a hospital or to the general practitioner (GP) on call. In Flanders, the GPs often organise this on call service for a specified region in a central location called General Practice Cooperatives (GPC, “Wachtpost” in Dutch). Patients do not know the characteristics of these different out of hours (OOH) services and find it hard to estimate the urgency of their own complaints. Therefore, many patients with presentations suitable for primary care go directly to the ED. This leads to additional costs for both government and patient, long waiting times at the ED and a high workload for the emergency physicians in Flanders.

**Aim of this project: we will advise you where to go**

The aim of this project is to deliver the most appropriate care for patients presenting at the ED. To achieve this objective a triage nurse will assess all patients presenting at an ED and advise them on the most appropriate point of care: GPC or ED. Triage is ‘the sorting out and classification of patients to determine priority of need’. Extended triage is triage combined with allocation to either ED or GPC, which is new in Flanders.

Many EDs in Flanders use the Manchester Triage system (MTS) in their regular care for triage within the ED, to sort patients according to the urgency of their presentation. We developed an extended MTS (eMTS) which foresees allocation to either ED or GPC. Scientific research is crucial to evaluate such triage systems before implementation is possible. Our study is a cluster-randomised trial. During one year, we will perform extended triage using the eMTS at one ED/GPC collaboration. Every four weekends, one weekend will serve as a control: the patient does not get an allocation advice.

At the end of this trial, we will deliver the first validated tool to perform extended triage and referral to primary care during OOH. Afterwards we aim to implement this tool throughout Flanders. Ultimately, we want to achieve a more efficient delivery of OOH care for Flemish patients with a cost reduction for the Flemish healthcare system.

**Scientific methodology of this project: is this service at this time the most appropriate for me?**

Three aspects determine the most appropriate OOH service for a specific patient at a specific time:

- *Medical aspects*: is this service capable of handling the medical problem in an efficient and qualitative way? Is it safe?
- *Financial aspects:* is it the most cost effective service for both government and patient?
- *Process aspects:* can the delivered service satisfy the perceived needs of patients and staff? What are the major barriers and facilitators? What can we learn from reported incidents? How can we improve safety?

This protocol contains a general introduction to the TRIAGE-trial and details on the medical aspect. For the two other aspects, we have separate research protocols.

We are using the MRC Framework for the Development and Evaluation of RCTs for Complex Interventions to Improve Health as a guidance for our trial(1). This framework consists of four steps:

- Developing: we have made a new eMTS tool before the start of this trial
- Piloting: we did perform a pilot study
- Evaluating: this is the concern of this application
- Reporting and implementing part of our utilisation strategy.

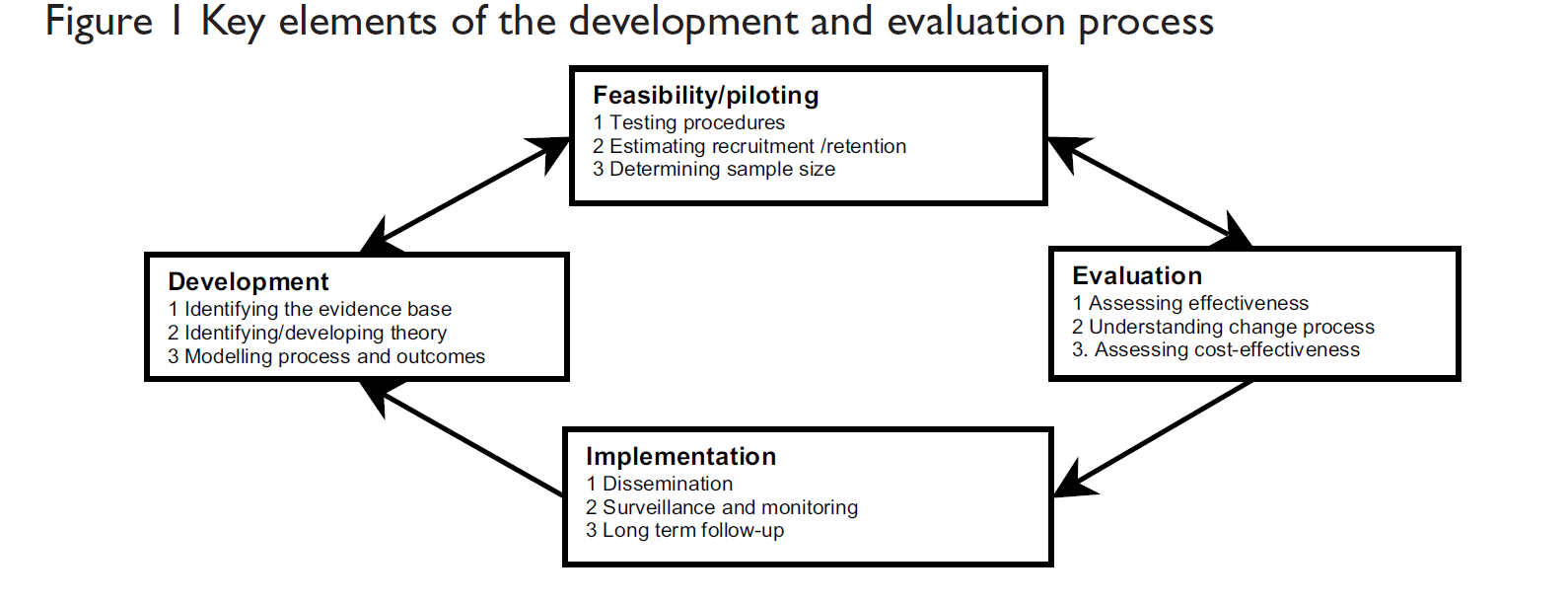

Figure 1: MRC framework: key elements of the development and evaluation process

###### Background

The aging society, increasingly demanding patients and cost pressures have steered most recent health care reforms in developed countries. The objective of improving health system efficiency through cost reduction while at the same time increasing patient and healthcare professional satisfaction and quality of care is the focus of these transformations and of this trial.

With regard to OOH situations, there is a trend to use hospital emergency departments (EDs) for standard and less urgent problems (2). At the same time, the decreasing number of GPs and their concerns about OOH workload and 24 h availability have led to an increasing workload and dissatisfaction. Policy makers have attempted to address these trends by organizing OOH care in a more structured and feasible way (3, 4). Ultimately, this led to a stronger centralisation of primary care centres. In Flanders, GPs run these centralised services (General Practice Cooperatives, GPCs) by themselves. In Flanders, the patient has unrestricted access to GPC and ED. The rise of GPCs did not reduce the use of nearby EDs, both services experienced a rise of patients (5). Patients choose for an ED because they believe they need technical examinations or want specialist advice (6). Patients however cannot judge the urgency of their complaint and are often not aware of the existence of a GPC (4). Worldwide EDs see a gradual increase in caseload. Extended triage in adjacent emergency and primary care departments is one of the solutions for this problem (7). In this extended triage, a nurse informs the patient about the most appropriate service for a specific problem at a specific moment in time (allocation). Ideally, triage is combined with a system of delayed care: some patients do not get immediate care but need to wait for care during office hours. In telephone triage, patients call a hotline before going to an OOH service. The telephone operator advises an OOH service using local protocols. The vast majority of patients in Flanders does not call but just heads directly to the ED or the GPC. We therefore suggest extended triage at the ED as an addition: a nurse informs the patient about the most appropriate service for a specific problem at a specific moment in time. Many factors can be related to citizens’ help‐seeking behaviour: timing (e.g. season), personal factors (e.g. age, gender, previous experiences, beliefs about the severity and susceptibility of a health problem), social factors (e.g. income and insurance) and cultural factors (e.g. origin and religion) (8). The influence of these factors on the choice of OOH service in Belgium and on the compliance to a triage advice is yet unknown. This project is the first research project to study the effectiveness of extended triage during OOH care in Flanders. Worldwide it will be the first prospective randomised controlled trial about extended triage.

Resources are scarce, and therefore it is essential to look at both effectiveness and resource use when introducing new treatments or treatment paths. However, only a limited number of studies already compared costs of patients treated by GPs and ED during OOH. Moreover, the methodology of those studies is weak. Giesbers and Giesen for example made use of simulations only(9). Others simply compared the costs of patients treated by a GP or in the ED, without taking into account that patient characteristics might differ between the two settings (10, 11). We will collect expenditure data and make use of a cluster-randomized trial to get an unbiased estimate of the difference in expenditures due to the extended triage.

Process evaluation is vital for assessing how external factors influence the delivery and functioning of interventions. It can help to explain how an intervention is working in a specific context to change the behaviours of specific target groups. This is particularly important in trials of complex interventions in ‘real world’ organisational settings such as out of hours care. Process evaluation seeks to identify the facilitators and barriers in an environment that may support/hinder implementation of extended triage in Flanders.

###### Study sites and partners

The TRIAGE-trial is a collaboration between the University of Antwerp, Antwerp University Hospital, clinicians at a GPC (Antwerpen Oost) and the ED of a general hospital (AZ Monica Deurne). In 2016, the GPC moved to a location adjacent to the ED. In 2016 the GPC had about 10 000 consultations during the weekends (from Friday 7pm until Monday 7am) for a population of almost 150 000 inhabitants. All 110 GPs working in the surroundings of the GPC are obliged to work at the GPC on average one shift per month. The ED treated about 35 000 patients in 2016 of which about 8 000 during the weekends. It has a 24 hours service. About eight emergency physicians staff the ED. The ED closely collaborates with the Antwerp University Hospital. Registrars from this University Hospital work in the ED as assistant emergency physicians. The surrounding area is a mix of middle-income neighbourhoods and ethnically diverse deprived neighbourhoods. We have established a local clinical committee to represent the study site partners: the head of the GPC, the head of the ED, the head nurse of the ED, two GPs and two nurses. The idea of this trial comes from the clinicians of this site themselves. Consequently, their motivation to carry out this trial is high. Because we will include almost all patients at the ED during the weekend, one site is definitely enough to include sufficient patients. If we find positive results, it will be possible to carry out a large multi-centre implementation and monitoring for a lower effort/cost.

###### Proof of Concept

The Manchester Triage system (MTS) is a validated tool for prioritisation of treatment at the ED (17). Allocation of patients either to an ED or to primary care was not the goal of its development but became a new goal afterwards. The manual of MTS contains a methodology to make a local extension with allocation. Several studies have already assessed the suitability of the MTS for referral to primary care.

- In a small (n= 115) prospective observational trial in the Netherlands children within the two lowest urgency categories of the MTS were referred to primary care. The need for extensive treatment and hospitalisation was assessed. The authors concluded these patients were suitable for primary care referral with some exceptions (18).
- In a large (n=3129) observational study in the Netherlands patients were allocated to the ED according to the MTS using a similar eMTS. The authors concluded that low urgent self-referrals with the exception of extremity problems can be treated efficiently and safely by a GP (19).
- In a small (n=264) interventional trial in Brazil, patients with less urgent problems were diverted to primary care. The authors concluded that this diversion might be feasible and safe with reasonable patient satisfaction (20).

Although a proof of concept, this research is definitely not enough to implement the studied systems in Flanders. The only large trial was a non-randomised observational trial without a financial evaluation. The limited number of studies with an economic assessment, merely collect costs in the different settings (10) or make use of a hypothetical calculation (9). The studied healthcare systems differ significantly from the Flemish situation making it impossible to use the same eMTS.

###### Pilot Study

We conducted a pilot study to collect baseline data and to assess the feasibility of cooperation between ED and GP. Preceded by a pre-measurement of three months we conducted an awareness-raising campaign for patients during five months. The intervention consisted of general information about when to seek help at an ED or GPC during the weekend distributed through leaflets, posters and broadcasting on a screen in the waiting room. Before the intervention, 1.7% of the presenting patients went spontaneously to the GPC after entering the ED. During the intervention, this increased to 5.4%. The switching patients had a typical primary care profile. There were no safety incidents. The GPs referred 5.7% of these patients back to the ED, a rate similar to the overall GPC population and current literature. This pilot study proves patient shifting is feasible and safe. Collaboration (including data collection) between the researchers and clinicians went smoothly.

###### eMTS

The Manchester Triage System (MTS) is one of the most commonly used triage systems in Europe(21). It enables nurses to assign a clinical priority to patients, based on presenting signs and symptoms, without making any assumption about the underlying diagnosis. The MTS assigns patients to one out of five urgency categories, which determine the maximum time to first contact with a physician (22). In the TRIAGE-trial, we will use an extended MTS (eMTS): we did not change the MTS but added some extra questions to allow allocation to ED or GPC. Because of legal and practical reasons, we have decided to allow allocation to primary care only for the two lowest urgency categories. The patients in the green category (called standard urgency) have to see a doctor within two hours. For the blue category (called non-urgent) the maximum period is four hours. We followed a consensus procedure to develop this eMTS with at least two professionals of all involved disciplines. We started with an online survey about the role for the GP for each of the 52 presentational flow charts of the MTS. Afterwards we rolled out group discussions focussing on those flow charts not reaching a consensus online. In Figure 2, we demonstrate the eMTS flow chart for shortness of breath in children.

Figure 2: eMTS flow chart for shortness of breath in children. Available upon request.

##### Research Approach

###### A diagnostic Intervention

A nurse will see the patient ideally within ten minutes after arriving at the ED. The nurse will classify a patient’s presentation using the MTS, which will enable to make a categorisation of urgency. If the patient’s urgency category is either blue or green, the nurse will use the eMTS to assess suitability for primary care. This assessment depends on the presentational flow chart used: for some flowcharts, all or none of the blue/green presentations will be allocated to primary care, for others the nurse has to ask additional questions (discriminators). The result of this extended triage is a shared decision of the patient and the nurse: the patient is (or is not) eligible for primary care. Because not all possible questions and signs are incorporated in the eMTS the triage nurses will have the possibility to overrule the eMTS advice in both directions. An adapted registration system will help the nurse select and follow the correct flowchart and will give an eMTS advice once all questions have been assessed. Patients will have the right to refuse the advice of the nurse without any further consequences for their treatment at the ED. We will provide all participating nurses and physicians with a detailed manual in Dutch about the intervention and the research protocol.

###### Hypotheses and Research questions

**Medical aspect**

*Our primary hypothesis:* the extended triage intervention will lead to a substantial increase in the proportion of study patients seen by the GP*.*

Without the intervention, all study patients can be divided in three categories: seen by ED, seen by GPC and left without being seen (after triage). By adding the compliance of the patient to the allocation advice, we get the following table:

|  | Patient goes to GPC | Patient goes to ED | Left without being seen |
| --- | --- | --- | --- |
| Advice GPC | Compliant patient GPC | Non-compliant patient GPC | Left without being seen type 1 |
| Advice ED | Non-compliant patient ED | Compliant patient ED | Left without being seen type 2 |

Table 1: patient categories

The proportion of study patients seen at the GPC is a combination of patients compliant to a GPC advice and patients not compliant to an ED advice. The latter category will probably be small.

*Research question:*

1. What is de the difference in the proportion of study patients seen by the GP between intervention and control weekends?
   1.1. Is this proportion associated with the patient’s presentation (chosen eMTS flowchart, MTS urgency, physician’s diagnosis)?
   1.2. Is this proportion associated with the crowding of the ED and/or GPC?
   1.3. Is this proportion associated with the operating triage-nurses?
   1.4. Is this proportion associated with the patient’s background characteristics? (sex, age, community, insurance status)?
   1.5. Is there daytime or seasonal variation in this proportion?

*Secondary hypothesis:* a nurse using the eMTS is able to detect patients suitable for primary care.

The results of the triage consult is either a GPC or ED advice. Arbitrarily we will call a GPC advice a positive diagnosis. The gold standard is the opinion of the physician (either emergency physician or GP) after the consultation. This standard is used in almost all of the literature(23), as no better gold standard is available. We are aware of some disadvantages: this opinion depends on the personality and other characteristics of the treating physician and some patients have presentations suitable for both ED and GPC. On the other hand, agreement between nurse and physician is important for the feasibility of extended triage.

The way we will measure the correctness of the triage advice depends on whether the patient is compliant to this advice. This makes Table 1 more complex:

|  | Patient goes to GPC | | Patient goes to ED | | |
| --- | --- | --- | --- | --- | --- |
| Opinion physician* | GPC correct | Had to go to ED | ED correct | Had to go to GPC | |
| Advice GPC | Correct decision 1 | False positive 1 | False positive 2 | | Correct decision 2 |
| Advice ED | False negative 1 | Correct decision 3 | Correct decision 4 | | False negative 2 |

Table 2: patient categories for the eMTS as a diagnostic instrument.

Because our eMTS is risk adverse, we already know we will have a certain amount of false negative 2: patients who are compliant to the advice of ED although they had to go to the GPC according to the emergency physician. This category is not our primary focus.

We are mostly interested in false positive 1: patients who complied with the advice GPC but should have stayed at the ED. Some of these patients might be subject to a safety risk. For example, a patient with a heart attack allocated to the GPC might lose valuable time. This category is the same as the proportion of study patients allocated to the GP but referred back to the ED. We will give this category special attention in our process analysis.

Finally, we are also interested in the category correct decision 2: patients who are not compliant to a correct GPC advice. These patients made an inefficient decision so there is room for improvement.

*Research questions:*

2. What is the validity of the eMTS as a preliminary diagnostic instrument to detect a primary care patient?
   2.1. What is the proportion of correct extended triage decisions?
        2.1.1. At the GPC (method 1): correct allocation means measure by referral to any ED
        2.1.2. At the GPC (method 2): correct allocation as assessed by the urgency category as registered by the GP
        2.1.3. At the ED (method 1): correct allocation means admission
        2.1.4. At the ED (method 2): correct allocation as assessed by the opinion of the emergency physician
   2.2. What is the proportion of patients wrongly assigned to GPC (all false positives)?
   2.3. What is the proportion of patients allocated to the GP and admitted afterwards?
   2.4. What is the proportion of patients wrongly assigned to GPC who did go to the GPC (type 1 false positives)
3. What is the proportion of patients who are not compliant to a correct GPC advice (Correct decision 2).
   3.1. What are the determinants for this non-compliance? (same determinants as studied in question 1.1-1.4)
4. What is the proportion of patients for which the nurse gives another advice comparing to the eMTS?
   4.1. What are the determinants for giving another advice? (same determinants as studied in question 1.1-1.4)
5. Does the intervention influence the number of patients left without being seen before and after the triage consultation?
6. What is the shift in workload during the different phases of this study for all participating healthcare providers (GPs, triage nurses and ED physicians)?
7. Does the epidemiology in terms of reasons for encounter and diagnoses at both OOH services differ between intervention and control weekends?
8. What is the proportion of patients who return to the study ED within two weeks for each of the categories described in table 2?

##### Design of the clinical trial

**A controlled longitudinal prospective intervention trial**

To answer the above-mentioned research questions we need a longitudinal prospective intervention trial.

To allow definitive conclusions for our research questions we need a control group. Historical controls will have too much confounding (not all data is available, the ED was organised in a different way, the GPC was unknown for many patients). Individually randomised controls are impossible from a practical point of view and because of contamination (patients will talk to each other and will not understand why some get a GPC advice and others not). Finally, randomisation by shift is not possible because they overlap in time. Therefore, randomisation is only possible by weekend. We will roll out one control weekend out of every four intervention weekends.

During a control weekend, all data registration and collection will be the same as during intervention weekends but we will not inform patients about their allocation advice. The emergency physician will see all patients deciding to stay at the ED, without influence of the triage advice. As in standard clinical care, patients will have the right to change their mind and go spontaneously to the GPC. Another difference is the communication to the patient: during intervention weekends, we will inform them about the intervention using leaflets and broadcasting in the waiting room of the ED. During control weekends, we will only inform about MTS-triage in general but no about the GPC. We will always mention the iCAREdata opting-out procedure.

We have chosen to use three times more intervention than control weekends because of three reasons. Firstly, safety monitoring is not possible during control weekends (the eMTS is risk adverse, we do not expect safety issues to occur on a weekly basis). Secondly, some of the research questions (2, 3 and 4) mainly concern the intervention weekends. Finally, we need sufficient patients to have enough data for every subgroup we will study during the intervention weekends (see Table 2).

**Inclusion and Exclusion**

We will systematically asses **all patients presenting at the ED during the weekend** for inclusion. The GPC is only open during the weekend starting from Friday 7 pm until Monday 7 am. The study inclusion will stop at 6 am to make sure the GP can leave on time. By presentation at the ED we mean the patient is registered by the receptionist of the ED as a patient. Patients who just pass the ED to enter the hospital or who go directly to a medical department (e.g. obstetrics) are thus not considered for inclusion. The availability of a Belgian citizen national insurance number is the only inclusion criterion. This number is necessary to send the data needed for this study to the iCAREdata database (see below). For the vast majority of patients this number is available. It will not be available for patients who present for the first time at the hospital and forgot all official documents. For patients without a Belgian identity (foreign citizens without Belgian health insurance, people without a legal status and babies less than one week old) the number is never available. The EDs own software allows us to calculate the number of excluded patients.

Patients arriving at the ED by an ambulance with a doctor or nurse (in Dutch called MUG or PIT) is the only exclusion criterion (these patients have already passed through a sort of triage). We estimate an exclusion rate of 15 patients per weekend.

**Sample size**

For this trial, we consulted our subcontractor StatUA (core statistical facility of the University of Antwerp). If we only look at our primary research question (“What is the difference in the proportion of patients presenting at the ED and seen by the GP between intervention and control weekends”), straightforward power computations will tell that two weekends (one with, and one without intervention) are sufficient to provide empirical evidence of a statistically significant shift. However, as we want to know the determinants of this proportion and perform safety monitoring (including incident analysis), we need observations over a longer period of time. We do not know the impact size of the determinants that might act as confounders in this context. Because the eMTS is risk adverse, safety issues will be rare and a long study period is required to identify and study possible safety issues. Finally, we expect nurses will need to build up experience with the eMTS before optimal results can be expected. For these reasons, we have chosen to include a large sample of patients by studying all weekends during one calendar year.

**Randomisation procedure**

We will assign the control weekends using randomisation software, taking into account possible confounders like bank holidays, school holidays, seasonal fluctuations…). We will not be able to blind the participants, the staff members nor the researchers. We will inform the staff about the nature of the upcoming weekend on Friday morning. Doing so, we aim to prevent anticipation of the staff (e.g. switching shifts or using less staff) and thus allocation bias. There is partial concealment of allocation for the patients as they do not know before their consultation whether it is a control or intervention weekend.

**Recruitment strategy and expected dropout**

Unlike most trials, we do not foresee any recruitment problems. In 2016, 8000 patients presented themselves during the weekend at the ED. There is a gradual increase in this number every year. Only patients participating in the process analysis will have to sign an informed consent. There will be an opting out procedure for all other patients. The ethical committee already waived informed consent for iCAREdata.

For each patient the study period is the time between presentation at the ED and discharge at either the ED or the GPC. This is a time span of only a few hours so the dropout rate will be negligible. Only when a patient leaves the site before consultation at the ED or GPC he or she is a dropout. This number of patients leaving the ED without being seen after triage (i.e. dropouts) is one of the safety indicators we will study. The number of these patients is currently less than five per weekend. We are able to measure this number using the triage reports through iCAREdata. Patients who leave without being seen before the triage-consultation will not be analysed.

Finally, we have the risk of overcrowding at the GPC. Currently this is not a big issue: in 2017, the longest waiting time was two hours but the vast majority of patients was seen within an hour after entering the GPC. If however we are to allocate a large part of the ED patients to the GPC this might change. A waiting time of over two hours at the GPC cannot be accepted. As soon as the number of patients waiting at the GPC reaches ten, the GPC will call in an additional GP. One hour after arrival of this GP, we will count the number of waiting patients again. If this number exceeds nine again, we will have to start the alarm procedure. The receptionist of the GPC will inform the ED that during at least one hour, allocation to the GPC is not possible. If after an hour, the number of patients in the waiting room is below 8, the procedure will stop.

##### Methodology for the medical aspect

**iCAREdata**

We have a large database about OOH care called iCAREdata (Improving Care And Research Electronic Data Trust Antwerp). The main aim of this project is to develop a central, clinical research database in out-of-hours (OOH) care in Flanders. It contains routine data of 1 127 000 patients and 9 GPCs. We develop this database since 2014 in sync with the most recent legal, ethical and privacy aspects present in Belgium and Europe. It contains data of patients using OOH care in a large part of Flanders starting from January 2015 to the present (15, 16). One crucial aspect of the project is the unique way it links data between different health care services using a coded national number. Subsequently, we are able to study the chain of care that patients follow in OOH care. This gives a broader view on what is exactly happening with patients suffering an unplanned medical problem. Because of the anonymity, we can answer most of the research questions in this application without the necessity of an individual informed consent.

**Data collection**

For more details on this subject we refer to our data collection plan. The data necessary for this work package are part of a routine clinical report. We will use three types of these reports: triage-consultation reports, ED consultation reports and GPC consultation reports. We will collect these data using the iCAREdata database. This database can link our three types of clinical reports using an irreversibly coded national number. Due to data protection rules, the iCAREdata data manager will execute all queries and only aggregated results will be transferred to the researchers. For more complex statistical analysis, the data manager will transfer an analysis specific table or the analysis will be carried out on the computer of the data manager in cooperation between the researcher and the data-manager. A detailed data management plan is available.

| Research question | Variable collected | Data source |
| --- | --- | --- |
| Number of exclusions | Number of presenting patients within study time (Friday 19.00-Monday 6.00) without Belgian national number. | ED-software (E.care) |
|  | Number of patients arriving in ambulance with nurse or doctor | iCAREdata: triage-consultations:”ContWijze” = “MUG” or “PIT” |
|  | Number of patients opting out | Manual recording: staff will contact the researchers to opt out specific patients. |
| Characteristics of excluded patients | To be defined: demographics | E.care |
| 1 | Numerator: Number of patients with a primary care advice after extended triage | iCARE data: number of triage-consultations with “ContVerwerk” = “wachtpost_patOK” or “wachtpost_patNotOK” and a GPC contact in iCAREdata |
|  | Denominator: Number of study patients | iCARE data: number of included triage-consultations |
| 1.1 | eMTS flowchart and category chosen by the nurse | iCAREdata: triage-consultations: flowchart = “DiagnTekst”, “DiagnCod”, urgency category = “ContUrgentie”  iCAREdata: ED-consultation: DiagnTekst, DiagnCod |
| 1.2 | Number of patients presenting at the ED and GPC in the hour before the start of a triage consultation | iCAREdata: triage-consultations (total number of patients seen at the ED) iCAREdata: triage-consultations (timestamp: start of triage consultation) iCAREdata: GP-consutations (all, not only included patients) |
|  | Subjective experience of workload by nurse (Likert-scale) | ContArbeidsongeschikt |
| 1.3 | Coded registry number or pseudo-number of the triage nurse | iCAREdata: triage-consultation= “IdArts” |
| 1.4 | Coded national number, sex, age, community (ZIP code), insurance status (higher reimbursement yes or no) of the patient | iCAREdata: triage-consultation is linked to a table with patients with these characteristics. |
| 1.5 | Time of start triage consultation | iCAREdata: triage-consultations (timestamp: start of triage consultation) |
| 2/3 | At GPC: number of patients referred to the ED  At ED: numer of patients with property “Had to go to GPC” as filled-in by the emergency physician | iCAREdata: GP-consultation: ContUrgentie, ContVerwerk  iCARE data ED-consultation: ContUrgentie, ContVerwerk, |
|  | Referral (if this variable contains “to ED”) |  |
|  | See 1.1-1.4 |  |
| 4 | eMTS flowchart and category chosen by the nurse | See 1.1 |
|  | Advice given by the nurse | iCARE data triage-consultation: “ContVerwerk” |
| 5 | Number of patients left before triage = no triage nor ED report available | Directly for E.care: a number per weekend, no patient details |
|  | Number of patients left after triage = Availability of a GP or ED report after triage consultation | iCAREdata: triage, GP and ED |
| 6 | Total number of patients seen in one shift (determined by a series of reports with the same coded registry number) | iCARE data: triage, GP and ED: ArtsId  For GPC: also for not included patients. |
| 7 | Reasons for encounter and diagnosis using the Belgian thesaurus and ICPC-2/ICD-10 | iCAREdata: GP and ED consutlations: “DiagnCod”; “ContactRedenCod” |
| 8 | Availability of a report | iCARE data: ED |

**Data analysis plan**

For more details we refer to our data analysis plan. The parameters of interest in questions 1 through 8 are (conditional) proportions, which calls for analysis by a non-linear model (e.g. binary logistic regression). The study design results in the creation of nested data, where patients are nested in weekends, and weekends are nested in (multiple) nurses. Appropriate analysis of such auto-correlated data requires a nonlinear mixed model. During the first months, data will be used to produce point-estimates of these proportions (e.g., patients seen by GP, correct triage decisions, all false positives, type 1 false positives, patients left without being seen, etc.). With increasing duration of the study, incoming data will be used to explain variability of these probabilities in terms of patient background characteristics, operating nurses/emergency physician, season, etc. Statistical assistance will be called for in order to conduct these analyses. We refer to our data analysis plan for more details.

###### Time Line

Prepartory Phase 1/10/2018-01/01/2019

Introduction phase 02/01/2019-31/01/2019

Interim Analysis 1/2/2019 28/2/2019

RCT phase 1/3/2019-31/12/2019

Analysis 1/1/2020-31/9/2021

###### Attachment : World Health Organization Trial Registration Data Set

| **Data category** | **Information** |
| --- | --- |
| Primary registry and trial identifying number | ClinicalTrials.gov |
| Date of registration in primary registry | Not yet registred |
| Secondary identifying numbers | N/A |
| Source(s) of monetary or material support | Research Foundation - Flanders (FWO). Grant Number T000718N. |
| Primary sponsor | Research Foundation - Flanders (FWO). |
| Secondary sponsor(s) | None |
| Contact for public queries | Veronique Verhoeven, |
| Contact for scientific queries | Hilde Philips, |
| Public title | the TRIAGE-trial: Triaging and Referring In Adjacent General and Emergency departments |
| Scientific title | Triaging and Referring In Adjacent General and Emergency departments (the TRIAGE-trial): a cluster randomised controlled trial |
| Countries of recruitment | Belgium |
| Health condition(s) or problem(s) studied | Any problem for which a patient might seek ED help. |
| Intervention(s) | Nurse led triage and when applicable referral to a General Practice Cooperative |
| Key inclusion and exclusion criteria | Inclusion: all patients presenting at the ED during the weekend.  Exclusion:   - No Belgian citizen national insurance number available - Arrived at the ED by ambulance with doctor or nurse |
| Study type | Interventional, cluster randomized Clusters: weekends Allocation: randomized Masking: None Primary purpose: healthcare improvement |
| Date of first enrolment | January 2019 |
| Target sample size | N/A, one year (2019) |
| Recruitment status | Recruiting |
| Primary outcome(s) | proportion of study patients seen by the GP |
| Key secondary outcomes | Association of primary outcome with:   - patient’s presentation (chosen eMTS flowchart and urgency)? - occupancy rate of the ED and/or GPC - operating triage-nurses and/or the operating emergency physician - patient’s background characteristics? (sex, age, community, insurance status) - seasonal variation   Correctness of the nurse’s allocation advice with the treating physician’s opinion as the gold standard  Changes in the expenditures for the Health Insurance System (HIS) and the patient and its determinants |

Attachment 2: future CONSORT flow-chart

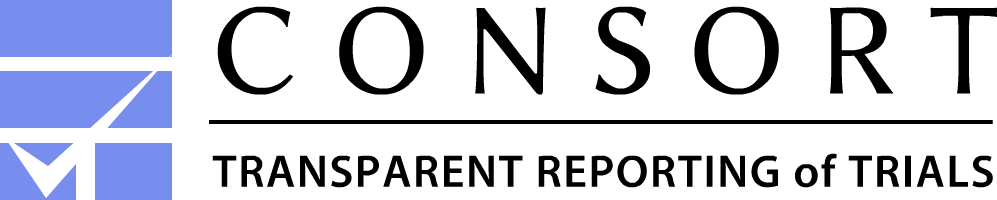

**CONSORT 2010 Flow Diagram**

Follow-Up

Analysed (n= )
♦ Excluded from analysis (give reasons) (n= )

Analysis

Lost to follow-up: only possibility is left without being seen before or after triage

Lost to follow-up: only possibility is left without being seen before or after triage (type 1 and 2)

Allocated to interventionweekends (n= )

♦ Received allocated intervention (n= )

♦ Did not receive allocated intervention (Left without being seen before triage)

Allocated to control weekends (n= )

Analysed (n= )
♦ Excluded from analysis (give reasons) (n= )

Excluded (n=	)

♦  No Belgian citizen national number (n= )

♦  Declined to participate, opting out (n= )

♦  Arriving by an ambulance with doctor or nurse

Enrollment

Assessed for eligibility (n= )

All patients getting triage

Randomized (n= )

Allocation

Declaration of interests

Stefan Morreel is working as a GP in the surroundings of the research site. He has obligatory on call duties at the GPC for which he receives honoraria. He is board member of the GPC for which he receives attendance fees. He was paid by the iCAREdata project during five months in 2018. He does not have any paid consultancies with pharmaceutical or other companies and has not received lecture fees for lectures given at international conferences.

Veronique Verhoeven received a travel fee for a presentation at the Yakult probiotics conference in Tokyo in 2009 and in London in 2013. Both events are unrelated to the current trial.
She received funding from Tilman n.v., a food supplement company, for an independent investigation of the cholesterol lowering effect of a red yeast rice food supplement, in 2015. She does not have any paid consultancies with pharmaceutical or other companies. She has not received lecture fees for lectures given at international conferences.

Koen Monsieurs was principal investigator for the VPAC trial, a prospective observational clinical study sponsored by Zoll Medical Corporation to verify an algorithm used to predict cardiopulmonary events in patients presenting to the emergency department. He does not have any paid consultancies with pharmaceutical or other companies. He has not received lecture fees for lectures given at international conferences.

Hilde Philips was working as a GP in the surroundings of the research site until 2017. She has no further connections to the board of the GPC, nor the Emergency Department of the adjacent hospital. She was principal investigator in several trials at GPCs in Flanders (Turnhout, Antwerpen-Noord). She was one of the investigators of the validation of 1733 protocols (telephone triage), commissioned by the health government of Belgium. She is coördinator of the iCAREdata project. The iCAREdata project will be used for data-collection in the current study. Both projects are independently financed and have a different scope. She did not receive lecture fees for lectures given at international conferences.

Diana De Graeve is an economist doing research on health topics. Research includes economic evaluations of drugs or medical devices. She occasionally had paid consultancies for firms or received lecture fees for lectures at conferences related to these economic evaluation studies or to the role of economics in the health care sector. Consultancy fees and lecture fees were not related to ED or GPC use in general or triage in particular.

### Minor changes to the study protocol after trial registration

#### Outcome measures

- The description of the primary and secondary outcomes have been modified to make the report more readable without changing their meaning
- In the original study protocol (but not in the trial registration!) the objectives were described as hypothesis. The authors have purposely chosen to register and report the trial as objective driven in stead of hypothesis driven as these hypothesis were not clearly enough defined and not always in line with the used methodology
- The terminology was slightly changed: the first ‘other outcome’ measure was reformulated and because of its importance renamed as the ‘secondary outcome’. All remaining secondary and other outcomes were renamed as ‘additional outcomes’.
- Association between the primary outcome and presenting complaint expressed as the title of a Manchester Triage System presentation: reporting about 53 different categories is unreadable and raises statistical issues because some categories were seldomly chosen. We have re-categorised them into 15 clinically relevant categories. (see supplementary table 4)
- Association between the primary outcome and subjective workload at the ED as judged by the emergency nurse: the number of categories of this variable has been reduced from four to three because “uncontrollable busy” was rarely chosen.
- Association between the primary outcome and subjective workload at the GPC: this variable was ultimately not available to the researchers as they were only allowed to collect data from the included patients.
- Association between the primary outcome and the age of the patient: this variable was categorised into age intervals to make the report more readable/consistent
- Association between the primary outcome and the ZIP-code of the patient: to make this variable more relevant to an international audience, it was dichotomised into nearby (communities surrounding the ED and covered by the GPC) or not nearby (all other communities)
- Association between the primary outcome and the season: there was a correlation with the primary outcome: winter versus other seasons (OR 1.47, 95%CI 1.23 to 1.92) but the only clusters during winter were Christmas, new year’s evening and the first three weekends of the trial making this finding prone to bias. There was no difference among the other seasons. To simplify the report this variable has been left out.
- Association between the primary outcome and the hour of the day of the patient's presentation: the analysis of this variable with 24 categories was hard to interpret and has been replaced by a more clinically relevant division: day, evening and night.
- ED Physician’s opinion on the ideal allocation of the patient: these physician’s had four options: ED, both ED and GPC, GPC or delay of care. Because delay of care is currently not allowed in Belgium and because the authors wanted to report predictive values, this variable has been dichotomised.
- Number of patients who returned to the ED within two weeks (in the protocol, not in the registration): this variable was ultimately not available to the researchers
- Sensitivity and specificity: positive and negative likelihood ratios were reported instead, see the statistical analysis plan for details
- The primary outcome after the trial ended was added as an additional outcome.

#### Eligibility Criteria

- Although always the intention, the exclusion of referred patients was not explicitly mentioned as an exclusion criterium (flaw in the study protocol).

### Supplementary Table 4: presentational flow chart categories

| Diarrhoea and vomiting | Abdominal complaints |
| --- | --- |
| Abdominal pain in adults | Abdominal complaints |
| Gastro-intestinal bleeding | Abdominal complaints |
| Neck Pain | Back and neck pain |
| Back Pain | Back and neck pain |
| Chest pain | Chest pain |
| Worried parent | Children |
| Abdominal pain in children | Children |
| Crying baby | Children |
| Shortness of breath in children | Children |
| Limping Child | Children |
| Abused or neglected child | Children |
| Unwell child | Children |
| Unwell baby | Children |
| Unwell newborn | Children |
| Eye problems | Eye problems |
| Limb Problems | Limb Problems |
| Apparently drunk | Mental complaints |
| Mental illness | Mental complaints |
| Behaving strangely | Mental complaints |
| Headache | Neurological complaints |
| Fits | Neurological complaints |
| Facial problems | ORL complaints |
| Dental problems | ORL complaints |
| Sore throat | ORL complaints |
| Ear problems | ORL complaints |
| Palpitations | Others |
| Abscesses and local infections | Others |
| Allergy | Others |
| Diabetes | Others |
| Rashes | Others |
| Asthma | Respiratory complaints |
| Shortness of breath in adults | Respiratory complaints |
| Bites and stings | Trauma and accidents |
| Chemical exposure | Trauma and accidents |
| Falls | Trauma and accidents |
| Head injury | Trauma and accidents |
| Overdose and poisoning | Trauma and accidents |
| Chest injury | Trauma and accidents |
| Assault | Trauma and accidents |
| Major incident | Trauma and accidents |
| Collapse | Unwell Adult |
| Unwell adult | Unwell Adult |
| Sexually acquired infection | Urinary or gynaecological complaints |
| Testicular pain | Urinary or gynaecological complaints |
| Urinary problems | Urinary or gynaecological complaints |
| Per Vaginam bleeding | Urinary or gynaecological complaints |
| Pregnancy | Urinary or gynaecological complaints |
| Burns and scalds | Wounds |
| Foreign body | Wounds |
| Wounds | Wounds |

ORL: Otorhinolaryngology
